# Multi-ancestry study of the genetics of problematic alcohol use in >1 million individuals

**DOI:** 10.1101/2023.01.24.23284960

**Authors:** Hang Zhou, Rachel L. Kember, Joseph D. Deak, Heng Xu, Sylvanus Toikumo, Kai Yuan, Penelope A. Lind, Leila Farajzadeh, Lu Wang, Alexander S. Hatoum, Jessica Johnson, Hyunjoon Lee, Travis T. Mallard, Jiayi Xu, Keira J.A. Johnston, Emma C. Johnson, Marco Galimberti, Cecilia Dao, Daniel F. Levey, Cassie Overstreet, Enda M. Byrne, Nathan A. Gillespie, Scott Gordon, Ian B. Hickie, John B. Whitfield, Ke Xu, Hongyu Zhao, Laura M. Huckins, Lea K. Davis, Sandra Sanchez-Roige, Pamela A. F. Madden, Andrew C. Heath, Sarah E. Medland, Nicholas G. Martin, Tian Ge, Jordan W. Smoller, David M. Hougaard, Anders D. Børglum, Ditte Demontis, John H. Krystal, J. Michael Gaziano, Howard J. Edenberg, Arpana Agrawal, Million Veteran Program, Amy C. Justice, Murray B. Stein, Henry R. Kranzler, Joel Gelernter

## Abstract

Problematic alcohol use (PAU) is a leading cause of death and disability worldwide. To improve our understanding of the genetics of PAU, we conducted a large cross-ancestry meta-analysis of PAU in 1,079,947 individuals. We observed a high degree of cross-ancestral similarity in the genetic architecture of PAU and identified 110 independent risk variants in within- and cross-ancestry analyses. Cross-ancestry fine-mapping improved the identification of likely causal variants. Prioritizing genes through gene expression and/or chromatin interaction in brain tissues identified multiple genes associated with PAU. We identified existing medications for potential pharmacological studies by drug repurposing analysis. Cross-ancestry polygenic risk scores (PRS) showed better performance in independent sample than single-ancestry PRS. Genetic correlations between PAU and other traits were observed in multiple ancestries, with other substance use traits having the highest correlations. The analysis of diverse ancestries contributed significantly to the findings, and fills an important gap in the literature.

## Introduction

Alcohol use disorder (AUD) is a chronic relapsing disease associated with a host of adverse medical, psychiatric, and social consequences^1^. Given observed heritability (*h*^2^∼50%^2^), there has been substantial progress made in genome-wide association studies (GWAS) of AUD and problematic drinking^3–9^, and also for measures of alcohol consumption^10,11^. A prior GWAS of problematic alcohol use (PAU, a phenotype based on a meta-analysis of highly genetically correlated (genetic correlations >0.7) AUD^7^, alcohol dependence [AD]^5^, and AUDIT-P [Alcohol Use Disorders Identification Test-Problem score, a measure of problematic drinking]^4,12^, N=435,563) identified 29□independent risk variants, predominantly in European (EUR) ancestry subjects^6^. Consistent with genetic studies of other complex traits, and the high polygenicity of PAU, larger and more ancestrally-representative samples need to be examined to outline the genetic architecture of these alcohol use traits.

A key finding from recent studies is that both AUD and AUDIT-P differ phenotypically and genetically from typical alcohol consumption^4,7^. AUD and AUDIT-P index aspects of disordered alcohol intake and correlate with genetic liability to negative psychiatric and psychosocial factors (e.g., higher major depressive disorder [MDD], lower educational attainment). An item-level study of the AUDIT questionnaire confirmed a two-factor structure at the genetic level, underscoring unique genetic influences on alcohol consumption and alcohol-related problems^13^ and noted that the genetics of drinking frequency were confounded by socio-economic status. A similar pattern – genetic distinctions between substance use disorder (SUD) vs. non-dependent use – has also been observed for cannabis use disorder and cannabis use^14^. Furthermore, aggregating across multiple substance use disorders suggests that problematic and disordered substance use has a unique genetic architecture that, while shared across SUDs, does not overlap fully with non-dependent substance use *per se*^15^.

Notwithstanding prior discovery of multiple genome-wide significant (GWS) loci for PAU, there are major gaps in our understanding of its genetic underpinnings. First, the estimated single-nucleotide polymorphism (SNP)-based heritability (*h*^2^) of AUD and PAU ranges from 5.6% to 10.0%^4–7^, reflecting substantial “missing heritability” compared to estimates based on genetic epidemiology, which show ∼50% heritability^2^. Second, most of the available samples used in human genetic studies – including for AUD – are of EUR ancestry; lack of ancestral diversity is a major problem both for understanding the genetics of these traits, and for potential applications of these genetic discoveries to global populations^16^. Our previous study in the Million Veteran Program (MVP) analyzed AUD in multiple ancestral groups^7^. However, non-EUR samples (N=72,387) were far smaller than EUR samples (N=202,004), resulting in inadequate statistical power and unbalanced gene discovery across ancestral backgrounds, which limits our understanding of the genetic architecture underlying the trait across populations.

To improve our understanding of the biology of PAU in multiple populations, we conducted substantially larger ancestry-specific GWAS of PAU followed by a cross-ancestry meta-analysis in 1,079,947 individuals from multiple cohorts. We identified 85 independent risk variants in EUR participants (almost tripling the number identified in previous studies) and 110 in the within-ancestry and cross-ancestry meta-analyses. We investigated the shared genetic architectures of PAU across different ancestries, performed fine-mapping for causal variants by combining information from multiple ancestries, and tested cross-ancestry polygenic risk score (PRS) associations with AUDIT-P in the UK Biobank (UKB) samples^17^. We combined genes identified by gene-based association analysis, transcriptome-wide association analysis (TWAS) and brain-chromatin interaction analysis, found dozens of genes linking to brain with convergent evidence. Drug repurposing analysis identified potential medications for further pharmacological studies, bringing forward the hope of novel biologically-directed medications strategies with the further potential of personalization. We conducted phenome-wide PRS analyses in biobanks from the PsycheMERGE Network^18^ in AFR and EUR-ancestry samples. We tested the genetic correlation between PAU and other traits, especially novel with respect to AFR samples where such analyses could not be conducted previously. These findings substantially augment the number of loci that contribute to risk of PAU, increasing power to investigate the causal relationships of PAU with other diseases, and identify novel druggable targets whose therapeutic potential requires empirical evaluation.

## Results

### Ancestrally diverse data collection

We collected newly genotyped subjects (most from MVP) and previously published data from multiple cohorts (MVP^19^, FinnGen^20^, UKB^17^, Psychiatric Genomics Consortium (PGC)^5^, iPSYCH^21,22^, Queensland Berghofer Medical Research Institute (QIMR) cohorts^23–25^, Yale-Penn 3^26^, and East Asian cohorts^27^) resulting in a total of 1,079,947 subjects (Table 1). Five ancestral groups were analyzed (Figure 1a): EUR (N=903,147), AFR (N=122,571), Latin American (LA, N=38,962), East Asian (EAS, N=13,551, all published in ref. 27), and South Asian (SAS, N=1,716). As in our previous study^6^, we utilized data on International Classification of Diseases (ICD)-diagnosed AUD (N_case_=136,182; N_control_=692,594), DSM-IV AD (N_case_=29,770; N_control_=70,282) and AUDIT-P (N=151,119), together defined as problematic alcohol use (PAU; based on high genetic correlations across these measures). The total number of AUD and AD cases was 165,952, almost double the 85,391 cases in the previously largest study^28^.

**Table 1.**
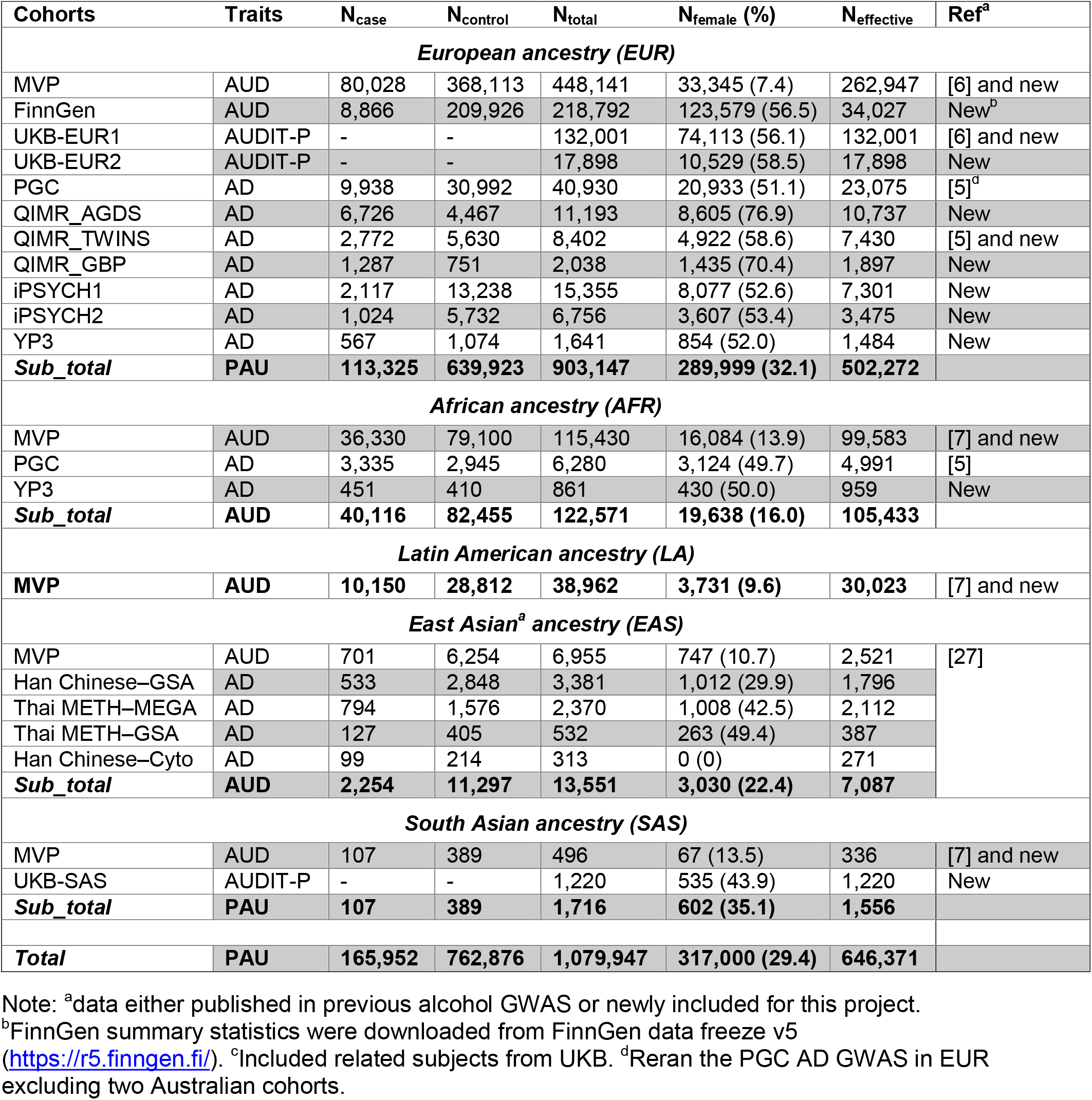
Demographics for cohorts in the meta-analysis of PAU. Cohorts are described in the Methods. UKB-EUR1: genetically defined White-British by UK Biobank; UKB-EUR2: genetically defined European non-White-British participants (see Methods); AGDS, the Australian Genetics of Depression Study; TWINS, the Australian twin-family study of alcohol use disorder; GBP, the Australian Genetics of Bipolar Disorder Study; iPSYCH1, phase 1 of iPSYCH; iPSYCH2, phase 2 of iPSYCH; YP3, Yale-Penn 3; N_effective_, effective sample size; Thai, study of the genetics of methamphetamine dependence in Thailand; GSA, Illumina Global Screening Array; MEGA, Illumina Multi-Ethnic Global Array; Cyto, Illumina Cyto12 array.

**Figure 1.**
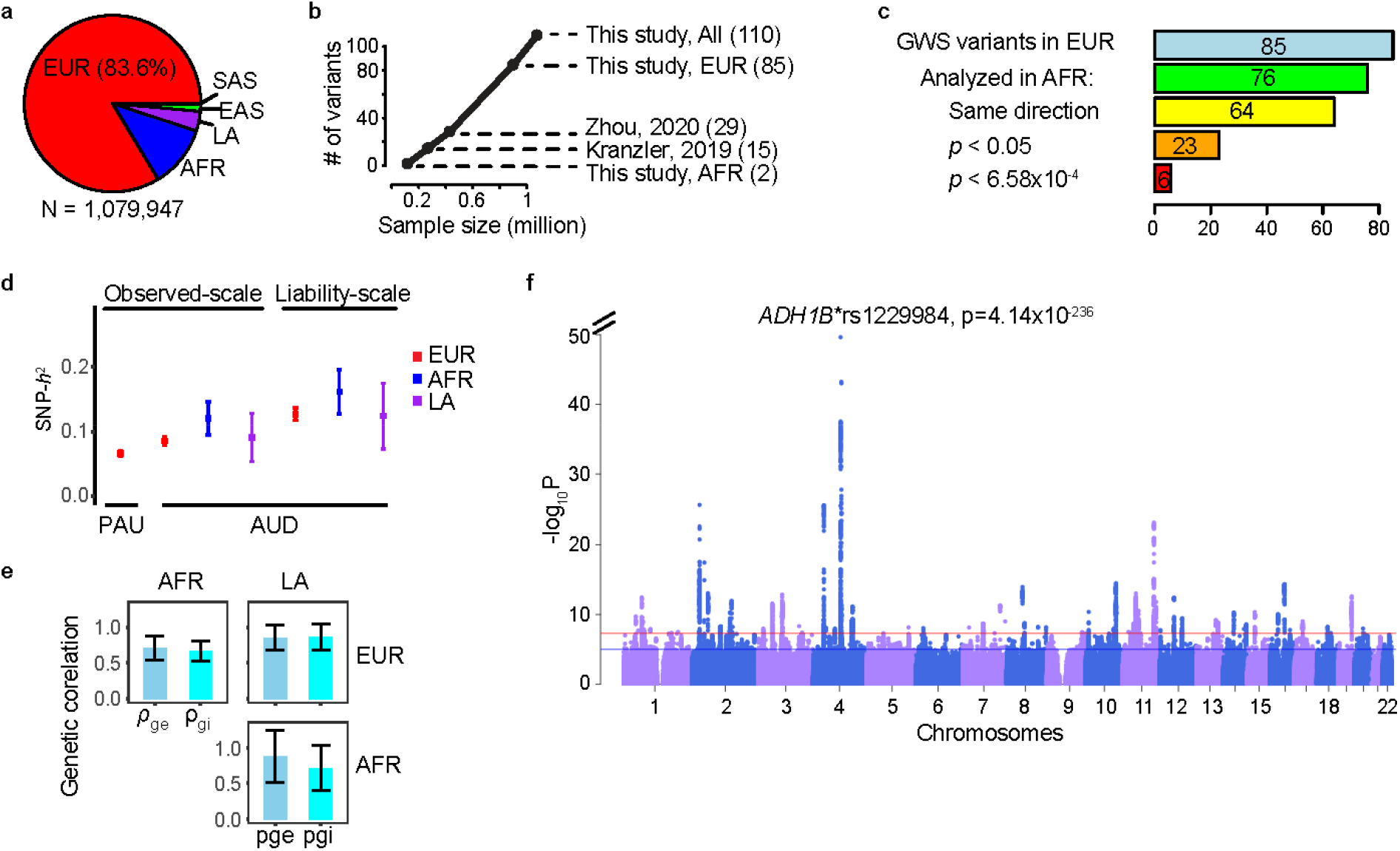
Genetic architecture of problematic alcohol use (PAU). **a**, Sample sizes in different ancestral groups. **b**, Relationship between sample size and number of independent variants identified. Kranzler et al., 2019: cross-ancestry meta-analysis for AUD; Zhou et al., 2020: PAU in EUR. **c**, Lookup for cross-ancestry replication in AFR for the 85 independent variants in EUR meta-analysis. Of the 85 variants, 76 could be analyzed in AFR (see Methods). Sign test was performed for the number of variants with same direction of effect (64/76, *p*=1.0×10^−9^). 23 variants were nominally significant in AFR and 6 were significant after multiple correction (*p*<0.05/76). **d**, Observed-scale and liability-scale SNP-based heritability (*h*^2^) in multiple ancestries. **e**, Cross-ancestry genetic-effect correlation (ρ_ge_) and genetic-impact correlation (ρ_gi_) between EUR, AFR and LA ancestries. Error bar is the 95% confidence interval. **f**, Genome-wide association results for PAU in the cross-ancestry meta-analysis. Red line is significance threshold of 5×10^−8^. EUR, European; AFR, African; LA, Latin American; EAS, East Asian; SAS, South Asian; GWS, genome-wide significant.

### Genome-wide association results for PAU

We performed GWAS and within-ancestry meta-analyses for PAU in five ancestral groups and then completed a cross-ancestry meta-analysis. In the EUR meta-analysis, 113,325 cases of AUD/AD, 639,923 controls and 149,899 participants with AUDIT-P scores were analyzed (Supplementary Figure 1a). After conditional analysis, 85 independent variants at 75 loci reached GWS (Methods, Supplementary Table 1) (see also Figure 1b). Of these variants, 41 are in protein-coding genes; of these, 5 are missense variants (*GCKR*\*rs1260326; *ADH1B*\*rs75967634; *ADH1B*\*rs1229984; *SCL39A8*\*rs13107325; *BDNF*\*rs6265).

Due to the smaller sample numbers, the non-EUR GWAS yielded fewer variants associated with PAU than did the EUR GWAS (Supplementary Table 1). The AFR meta-analysis found two independent *ADH1B* missense variants (rs1229984 and rs2066702) associated with AUD (Figure 1b, Supplementary Figure 1b); these were reported previously^7,26^. In the LA samples from MVP, only *ADH1B*\*rs1229984 (lead SNP) was identified (Supplementary Figure 1c). Two independent risk variants, *ADH1B*\*rs1229984 and *BRAP*\*rs3782886, were reported in EAS previously^27,29^. In the small SAS meta-analysis, one intergenic variant (rs12677811) was associated with AUD; however, this SNP was present only in the UKB (Supplementary Figure 1d).

Of the 85 lead variants identified in the EUR GWAS, 76 were either directly analyzed or had proxy variants in AFR (Methods, Supplementary Table 2, Figure 1c), 64 of which had the same direction of effect (sign test *p*=1.00×10^−9^). Of these, 23 were nominally associated (*p*<0.05) and 6 were significantly associated with AUD after multiple-testing correction (*p*<6.58×10^−4^). In LA, 15 of the EUR GWS variants were nominally significant (*p*<0.05) and 2 were significantly associated with AUD (rs12048727 and rs1229984). In EAS, 5 variants were nominally significant and two were significantly associated with AUD (rs1229984 and rs10032906). Only two variants were nominally associated with PAU in SAS (rs1229984 was not present in SAS).

We estimated the SNP-based heritability (*h*^2^) for PAU and AUD (excluding AUDIT-P from UKB) in EUR, AFR and LA; significant *h*^2^ estimates (range from 0.066 to 0.127) were observed (Figure 1d, Supplementary Table 3).

High genetic correlations were observed across the EUR, AFR, and LA ancestries (Figure 1e, Supplementary Table 4). The genetic-effect correlation (ρ_ge_) is 0.71 (SE=0.09, *p*=6.16×10^−17^) between EUR and AFR, 0.85 (SE=0.09, *p*=3.14×10^−22^) between EUR and LA, and 0.88 (SE=0.18, *p*=1.58×10^−6^) between AFR and LA. The genetic-impact correlation (ρ_gi_) is 0.67 (SE=0.07, *p*=2.78×10^−21^) between EUR and AFR, 0.86 (SE=0.09, *p*=3.52×10^−20^) between EUR and LA, and 0.72 (SE=0.16, *p*=9.63×10^−6^) between AFR and LA. The estimates involving smaller study populations were not robust (Bonferroni *p*>0.05).

In the cross-ancestry meta-analysis of all available datasets, we identified 100 independent variants at 90 loci (Figure 1f, Supplementary Table 1); 80 are novel findings for PAU. Of these, 53 variants were located in protein-coding genes, of which 9 are missense variants: *GCKR*\*rs1260326; *ADH1B*\*rs75967634, rs1229984, and rs2066702; *SCL39A8*\*rs13107325; *OPRM1*\*rs1799971; *SLC25A37*\*rs2942194; *BDNF*\*rs6265; and *BRAP*\*rs3782886. The cross-ancestry meta-analysis identified 24 more risk variants than the EUR meta-analysis, but 9 EUR variants fell below GWS (p-values ranging from 5.26×10^−6^ to 9.84×10^−8^). In total, 110 unique variants were associated with PAU in either the within-ancestry or cross-ancestry analyses (Figure 1b, Supplementary Table 1).

### Within- and cross-ancestry causal variant fine-mapping

We performed within-ancestry fine-mapping for the 85 clumped regions with independent lead variants in EUR (Supplementary Tables 5 and 6). A median number of 115 SNPs were included in each region to estimate the credible sets with 99% posterior inclusion probability (PIP) of causal variants. After fine-mapping, the median number of SNPs constituting the credible sets was reduced to 20. Among the 85 regions, there were 5 credible sets that include only a single variant with PIP ≥99% (presumably indicating successful identification of specific causal variants): rs1260326 in *GCKR*, rs472140 and rs1229984 in *ADH1B*, rs2699453 (intergenic), and rs2098112 (intergenic). Another 19 credible sets contained ≤5 variants (Figure 2a).

**Figure 2.**
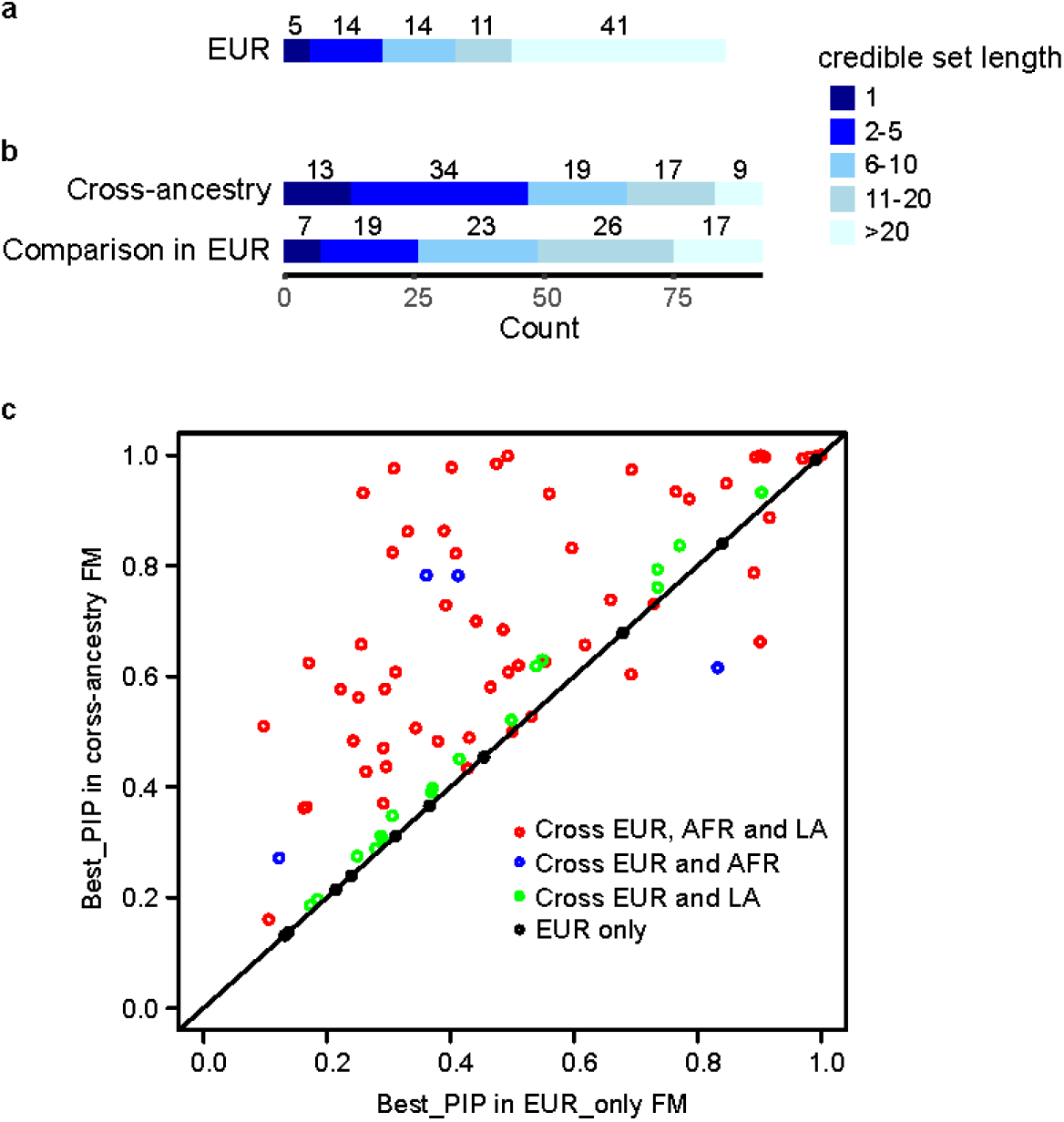
Fine-mapping for PAU. **a**, Fine-mapping of causal variants in 85 regions in EUR. **b**, 92 regions in cross-ancestry analysis were fine-mapped and a direct comparison was done for these regions in EUR. **c**, Comparison for the highest PIPs from cross-ancestry and EUR-only fine-mapping in the 92 regions. Red dots are the regions fine-mapped across EUR, AFR, and LA; blue dots are the regions fine-mapped across EUR and AFR; green dots are the regions fine-mapped across EUR and LA; black dots are the regions only fine-mapped in EUR. FM, fine-mapping.

We performed cross-ancestry fine-mapping to identify credible sets with 99% PIP for causal variants proximate to 92 independent lead variants in the cross-ancestry meta-analysis (Supplementary Tables 7 and 8). The median number of SNPs in the credible sets was 9. 13 credible sets contain only a single variant with PIP ≥99%; 47 credible sets contain ≤5 variants (Figure 2b). For example, fine-mapping the region proximate to lead SNP rs12354219 (which maps to *DYPD* on chromosome 1) identified rs7531138 as the most likely potential causal variant (PIP=48%), although this variant and rs12354219 (PIP=11%) are in high linkage disequilibrium (LD) in different populations (*r*^2^ ranges from 0.76 to 0.99). In a cross-ancestry meta-analysis rs7531138 showed significant association with schizophrenia (*p*=1.04×10^−8^), but rs12354219 (*p*=6.18×10^−8^) did not^30^ (although the two p-values were very similar). rs7531138 is also a lead SNP associated with educational attainment (*p*=1.74×10^−11^), unlike rs12354219 (*p*>5×10^−8^)^31^.

To compare within- and cross-ancestry fine-mapping, we performed fine-mapping for the above 92 regions using the same SNP sets and EUR-only LD information (Figure 2b & 2c). The median number of SNPs in the credible sets is 13, with 7 credible sets containing a single variant and 26 containing ≤5 variants, indicating that cross-ancestry fine-mapping improved causal variant identification, consistent with other studies reporting improved fine-mapping by including other ancestries^11^.

### Gene-based association analysis

We used MAGMA^32,33^ to perform gene-based association analyses. 130 genes in EUR, 9 in AFR and 6 in LA (for AFR and LA populations, all mapped to the ADH gene cluster), and 7 in EAS (mapped to either the ADH gene cluster or the *ALDH2* region^27^) were associated with PAU or AUD (Supplementary Table 9). There were no significant findings in SAS.

### Transcriptome-wide association analyses (TWAS)

We used S-PrediXcan^34^ to identify predicted gene expression associations with PAU in 13 brain tissues^35^. 426 significant gene-tissue associations were identified, representing 89 unique genes (Supplementary Table 10). Five genes showed associations with PAU in all available brain tissues, including *AMT* (Aminomethyltransferase), *YPEL3* (Yippee Like 3), *EVI2A* (Ecotropic Viral Integration Site 2A), *EVI2B* (Ecotropic Viral Integration Site 2B), and *CTA-223H9*.*9* (lncRNA). We also observed associations between PAU and the expression of alcohol dehydrogenase genes (*ADH1B* in the putamen (basal ganglia), *ADH1C* in 10 brain tissues, and *ADH5* in cerebellar hemisphere and cerebellum). Among the brain tissues, caudate (basal ganglia) had the most genes whose expression was associated with PAU (42 genes), followed by the putamen (basal ganglia) (39 genes). TWAS that integrated evidence across 13 brain tissues using S-MultiXcan^36^ to test joint effects of gene expression variation identified 121 genes (81 shared with S-PrediXcan) whose expression was associated with PAU (Supplementary Table 11).

### Linking risk genes to brain chromatin interaction

We used H-MAGMA^37^ to implicate risk genes associated with PAU by incorporating brain chromatin interaction profiles. 1,030 gene-chromatin associations were identified in 6 brain Hi-C annotations, representing 401 unique genes (Supplementary Table 12). 58 genes showed association with chromatin interaction in all 6 annotations, including *ADH1B, ADH1C, DRD2, EVI2A* and others that also showed evidence by TWAS in brain tissues.

### Convergent evidence linking association to brain

We examined overlapped genes by both gene-based association analysis and TWAS in brain tissues and/or H-MAGMA analysis using Hi-C brain annotations. Among the 130 genes associated with PAU in EUR, 60 were also implicated by TWAS findings either by single brain tissue (S-PrediXcan) or across brain tissues (S-MultiXcan), 82 have evidence of brain chromatin interaction, and 38 have evidence from both TWAS and Hi-C annotations including *ADH1B, DRD2, KLB* and others (Supplementary Table 9).

### Probabilistic fine-mapping of TWAS

We performed fine-mapping for TWAS using FOCUS^38^, a method that estimates credible gene sets predicted to include the causal gene that can be prioritized for functional assays. We detected 53 credible sets at a nominal confidence level (set at 90% PIP). These contained 145 gene-tissue associations with an average PIP of 32% (Supplementary Table 13). For the 19 gene-tissue associations having PIP >90%, 9 are from brain tissues (e.g., *ZNF184* expression in hypothalamus (PIP=0.94%), *MTCH2* expression in nucleus accumbens (basal ganglia) (PIP=99%), *SLC4A8* expression in dorsolateral prefrontal cortex (PIP=98%), *YPEL3* expression in cerebellum (PIP=100%), and *CHD9* expression in dorsolateral prefrontal cortex (PIP=100%).

### Drug repurposing

Independent genetic signals from the cross-ancestry meta-analysis were searched in OpenTargets.org^39^ for druggability and medication target status based on nearest genes. Among them, *OPRM1* implicated naltrexone and *GABRA4* implicated acamprosate, both current treatments for AUD. Additionally, the genes *DRD2, CACNA1C, DPYD, PDE4B, KLB, BRD3, NCAM1, FTOP*, and *MAPT*, were identified as druggable genes.

From the drug repurposing analysis using S-PrediXcan results, 287 compounds were significantly correlated with the transcriptional pattern associated with risk for PAU (Supplementary Table 14). Of these 287, 141 medications were anti-correlated with the transcriptional pattern. Of those, trichostatin-a (*p*=3.29×10^−35^), melperone (*p*=6.88×10^−11^), triflupromazine (*p*=7.37×10^−10^), spironolactone (*p*=2.45×10^−9^), amlodipine (*p*=1.42×10^−6^) and clomethiazole (*p*=1.30×10^−5^) reversed the transcriptional profile associated with increased PAU risk, targeted a gene near an independent significant locus in the cross-ancestry GWAS.

### Cross-ancestry polygenic risk score association

We tested the cross-ancestry PRS association with AUDIT-P in UKB using AUD summary data from EUR (leaving out the UKB AUDIT-P data), AFR, and LA. PRS-CSx was applied to calculate the posterior effect sizes for each SNP by leveraging LD diversity across discovery samples^40^. We validated the PRS associations with AUDIT-P in UKB-EUR2 and tested them in UKB-EUR1 (see Table 1). In the UKB-EUR1 samples, EUR-based AUD PRS is significantly associated with AUDIT-P (Z-score=11.6, *p*=3.14×10^−31^, ΔR^2^=0.11%). By incorporating GWAS data from multiple ancestries, the AUD PRS is more significantly associated with AUDIT-P and explains more variance (Z-score=13.6, *p*=2.44×10^−42^, ΔR^2^=0.15%) than the single ancestry AUD PRS.

### Genetic correlations

We confirmed significant positive genetic correlations (*r*_g_) in EUR between PAU and substance use and psychiatric traits^6^ (Supplementary Table 15). AD^5^ showed the highest correlation with PAU (*r*_g_=0.85, SE=0.07, *p*=4.49×10^−34^), followed by maximum habitual alcohol intake^9^ (*r*_g_=0.79, SE=0.03, *p*=1.24×10^−191^), opioid use disorder^41^ (*r*_g_=0.78, SE=0.04, *p*=1.20×10^−111^), drinks per week^11^ (*r*_g_=0.76, SE=0.02, *p*<1×10^−200^), smoking trajectory^42^ (*r*_g_=0.63, SE=0.02, *p*=2.47×10^−176^), and cannabis use disorder^14^ (*r*_g_=0.61, SE=0.04, *p*=4.85×10^−63^). We next tested *r*_g_ between AUD and 13 published traits with large GWAS in AFR (Figure 3, Supplementary Table 16). As in EUR, the traits with the strongest correlations were substance use traits. Maximum habitual alcohol intake^9^ (*r*_g_=0.67, SE=0.15, *p*=8.13×10^−6^) showed the highest correlation with AUD, followed by opioid use disorder^41^ (*r*_g_=0.62, SE=0.10, *p*=6.70×10^−10^), and smoking trajectory^42^ (*r*_g_=0.57, SE=0.08, *p*=3.64×10^−4^). Major depressive disorder^43^ and smoking initiation^11^ showed nominally significant (*p*<0.05) positive correlation with AUD and type 2 diabetes^44^ showed a nominally significant negative correlation.

**Figure 3.**
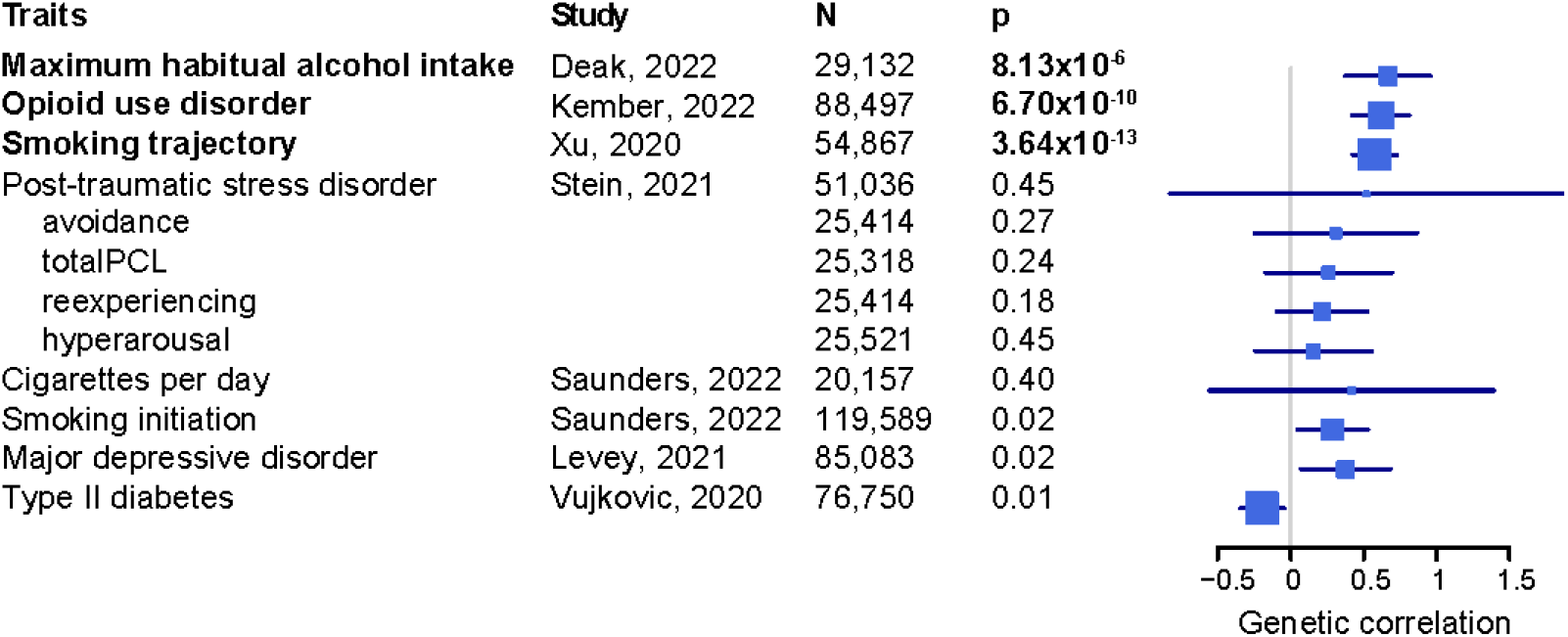
Genetic correlations between AUD and traits in AFR. totalPCL, total index of recent symptom severity by PTSD checklist for DSM-IV. Traits labeled in bold font are genetically correlated with AUD after Bonferroni correction (*p*<3.83×10^−3^). Error bar is 95% confidence interval.

### PRS for phenome-wide associations

We calculated PRS for PAU (based on the meta-analysis of PAU in EUR) in 131,500 individuals of European ancestry, PRS for AUD (based on the meta-analysis of AUD in AFR) in 27,494 individuals of African ancestry in 4 biobanks (Vanderbilt University Medical Center BioVU, Penn Medicine BioBank, Mount Sinai Icahn School of Medicine Bio*Me™*, and Mass General Brigham Biobank) from the PsycheMERGE Network, and conducted phenome-wide association studies (PheWAS). After Bonferroni correction, 58 of the 1,493 tested phenotypes were significantly associated with the PAU PRS in EUR, including 26 mental disorders, 8 respiratory traits, 5 neurological conditions, 4 infectious diseases, and 4 neoplasms (Supplementary Table 17, Supplementary Figure 2). For the 793 phenotypes tested in AFR, alcoholism (OR=1.25, SE=0.04, *p*=2.62×10^−7^), alcohol-related disorders (OR=1.21, SE=0.04, *p*=4.11×10^−7^), and tobacco use disorder (OR=1.09, SE=0.02, *p*=6.98×10^−6^) showed significant association with AUD PRS (Supplementary Table 18, Supplementary Figure 3).

We also conducted PheWAS in Yale-Penn, a deeply phenotyped cohort with comprehensive psychiatric assessments (substance use disorders [SUDs] and psychiatric disorders) and assessments for physical and psychosocial traits^26,45^. In EUR, the PRS of PAU was associated with 123 traits, including 26 in alcohol, 39 in opioid, 24 in cocaine and 17 in tobacco categories (Supplementary Table 19, Supplementary Figure 4), indicating high comorbidity and shared genetic components across SUDs. In AFR, the AUD PRS was associated with the DSM-5 AUD criterion count, alcohol-induced blackouts, frequency of alcohol use, and 3 individual AUD criteria: unsuccessful effort to decrease use, used more than intended, and continued use despite social/interpersonal problems (Supplementary Table 20, Supplementary Figure 5).

## Discussion

We report here the largest multi-ancestry GWAS for PAU to date, comprising over 1 million individuals and including 165,952 AUD/AD cases, more than double the largest previous study^28^. Considering the results from this study and previous GWAS, in all ancestral populations, we observed a nearly linear relationship between sample size and the number of risk variants discovered.

Convergent evidence supports substantial shared genetic architecture for PAU across multiple ancestries. First, of the 76 independent risk variants detected in EUR and represented in other populations, the majority have the same direction of effect in AFR (84.2%) and LA (81.6%). Twenty-three variants (30.3%) in AFR and 15 (19.7%) in LA were nominally replicated (*p*<=0.05), which is considerable given the appreciably lower sample size of these ancestral groups. Second, there are high cross-ancestry genetic correlations among EUR, AFR, and LA, ranging from 0.71 (between EUR and AFR) to 0.88 (between AFR and LA). Third, cross-ancestry meta-analysis substantially improved the power for gene discovery and resulted in the identification of 24 additional variants beyond the EUR-only results.

A total of 110 variants were associated with PAU in either within-ancestry or cross-ancestry analyses; of these, 9 are missense variants. These include rs1799971 in *OPRM1* which encodes the μ opioid receptor, which plays roles in regulating pain, reward, and addictive behaviors. This variant was also associated with opioid use disorder (OUD) in multiple large GWAS^41,46,47^. Previously, there were inconsistent candidate gene association results for *OPRM1*\*rs1799971 and AUD (reviewed in ref. 48). This is the first GWAS to confirm the association of rs1799971 in PAU; the risk allele is the same as for OUD. In contrast to an apparent EUR-specific effect of rs1799971 on OUD, the *OPRM1* association with PAU (*p*=6.16×10^−9^) was detected in the cross-ancestry meta-analysis. Further investigation in larger non-EUR samples is needed to assess the association of this SNP with SUDs in different population groups. Rs6265 in *BDNF* (brain-derived neurotrophic factor) encodes a member of the nerve growth factor family of proteins and has been investigated intensively in the past decades^49^; studies showed that this variant is associated with smoking traits^10^ and externalizing behavior^50^. Rs13107325 in *SLC39A8* (Solute Carrier Family 39 Member 8) has been associated with schizophrenia^51^, substance uses^6,7,10^ and many glycemic traits, and is critical for glycosylation pathways^52,53^.

Previous studies have shown that PAU is a brain-related trait with evidence of functional and heritability enrichment in multiple brain regions^6,7^. We performed gene-based association, TWAS in brain tissues, and H-MAGMA analysis in brain annotations. We identified 38 genes that were supported across multiple levels of analysis. For example, *ADH1B* expression in putamen was associated with PAU by TWAS, and with chromatin interaction in all 6 brain annotations by H-MAGMA, indicating additional potential biological mechanisms for the association of *ADH1B* with PAU risk through gene expression and/or chromatin interactions in brain, potentially independent of the well-known hepatic effect on alcohol metabolism. *DRD2* expression in cerebellar hemisphere and chromatin interaction in all brain annotations were also associated with PAU risk. Alcohol metabolism, as is well-reported, has affects that modulate alcohol’s aversive and reinforcing effects^54^, but also contributes to brain histone acetylation, gene expression and alcohol-related associative learning in mice^55^. The detailed molecular pathways and mechanisms involving changes in human brain need to be elucidated.

Independent genetic signals supported the two main AUD pharmacological treatments acamprosate and naltrexone: *GABRA4* is a target of acamprosate while *OPRM1* is a target for naltrexone. We identified genes known to be druggable; our multivariate analysis also provided evidence for several repurposable drugs. Trichostatin-a, a histone deacetylase inhibitor, showed effects on H3 and H4 acetylation and neuropeptide Y expression in the amygdala and prevented the development of alcohol withdrawal-related anxiety in rats^56^. Clinical trials showed that melperone, a dopamine and serotonin receptor antagonist, has inconsistent effects on alcoholic craving^57,58^. Spironolactone, a mineralocorticoid receptor antagonist, reduced alcohol use in both rats and humans in a recent study^59^. Clomethiazole, a GABA receptor antagonist, also showed effect of treatment for alcohol withdrawal syndrome^60^. Future clinical trials may use the evidence from this drug-repurposing analysis to prioritize drugs for further study.

PAU was positively genetically correlated with many psychiatric and substance use disorders and negatively with cognitive performance. Most of our genetic correlations with PAU, and all those in previous studies, were restricted to EUR populations, presumably because of insufficient statistical power in other populations. The PheWAS PRS also identified associations with medical phenotypes in EUR. With increasing number of AFR GWAS now published, mainly from MVP, we were able to estimate genetic correlations between AUD and a limited set of traits in AFR. As in EUR, AUD in AFR was genetically correlated with substance use traits including OUD, smoking trajectory (which identifies groups of individuals that follow a similar progression of smoking behavior), and maximum habitual alcohol intake. PheWAS of PRS in AFR from PsycheMERGE and Yale-Penn confirmed that AUD is genetically correlated with substance use traits. The lack of a wider set of phenotypes for comparison by ancestry is a continuing limitation.

Additional limitations include that the differences in ascertainment and phenotypic heterogeneity across cohorts might bias the results. Despite the high genetic correlation between AUD and AUDIT-P, they are not identical traits. Also, differences in ascertainment amongst the cohorts may have introduced additional biases; for example, considering the QIMR AGDS and GBP cohorts, the former have high major depression comorbidity, and the latter have high bipolar disorder comorbidity. (This heterogeneity would, however, have been more likely to limit discovery than to create false-positives.) Additionally, while we set out to include all available samples for problematic drinking in multiple ancestries, the sample sizes in the non-EUR ancestries were still small for gene discoveries and downstream analyses. The collection of substantial numbers of non-European subjects is a critical next step in this field.

In summary, we report here a large multi-ancestry GWAS and meta-analysis for PAU, in which we focused our analyses in three main directions. First, we demonstrated that there is substantial shared genetic architecture of PAU across multiple populations. Second, we analyzed gene prioritization for PAU using multiple approaches, including cross-ancestry fine-mapping, gene-based association, brain-tissue TWAS and fine-mapping, and H-MAGMA for chromatin interaction. We identified many genes associated with PAU with biological support, extending our understanding of the brain biology that substantially modifies PAU risk and expands opportunities for investigation using *in vitro* methods and animal models. These genes are potential actionable targets for downstream functional studies and possible targets of pharmacological intervention based on the drug repurposing results. Third, we investigated the genetic relationship between PAU and many traits, which for the first time was possible in AFR populations. Future increases in sample size will doubtless yield additional gains; this is particularly needed in non-EUR populations both for primary GWAS analyses and the analysis of other traits for comparison and to estimate pleiotropy.

## Methods

### Study design

In the previous PAU study^6^, the *r*_g_ between MVP AUD and PGC alcohol dependence (AD) was 0.98, which justified the meta-analysis of AUD (includes AUD and AD) across the two datasets; and the *r*_g_ between AUD and UKB AUDIT-P was 0.71, which justified the proxy-phenotype meta-analysis of PAU (including AUD, AD and AUDIT-P) across all datasets. In this study, we use the same definitions, defining AUD by meta-analyzing AUD and AD across all datasets, and defining PAU by meta-analyzing AUD, AD and AUDIT-P (Table 1).

### MVP dataset

MVP enrollment and genotyping have been described previously^19,61^. MVP is a biobank supported by the US Department of Veterans Affairs (VA) with rich phenotypic data collected using questionnaires and the VA electronic health record system (EHR). The Central VA Institutional Review Board (IRB) and site-specific IRBs approved the MVP study. All relevant ethical regulations for work with human subjects were followed in the conduct of the study and informed consent was obtained from all participants.

MVP genotype data were processed by the MVP Release 4 (R4) Data Team. 729,324 samples were genotyped using Affymetrix Axiom Biobank Array. Rigorous sample-level quality control (QC) served to remove samples with duplicates, call rates <98.5%, sex mismatches, >7 relatives, or excess heterozygosity. After QC, MVP R4 data contains 658,582 participants and 667,995 variants (pre-imputation). Pre-imputation QC removed variants with high missingness (>1.5%), that were monomorphic, or with Hardy-Weinberg Equilibrium (HWE) p-value ≤1×10^−6^, leaving 590,511 variants for imputation. As in our previous work^7^, we ran principal component analysis (PCA)^62,63^ for the R4 data and 1000 Genome phase3 reference panels^64^. The Euclidean distances between each MVP participant and the centers of the five reference ancestral groups were calculated using the first 10 PCs, with each participant assigned to the nearest reference ancestry. A second round PCA within each assigned ancestral group was performed and outliers with PC scores >6 standard deviations from the mean of any of the 10 PCs were removed. This two-stage approach resulted in the assignment of 468,869 European ancestry (EUR), 122,024 African ancestry (AFR), 41,662 Latin American (LA), 7,364 East Asian (EAS) and 536 South Asian (SAS) individuals for analysis.

Imputation was done by the MVP R4 Data Team. The entire cohort was pre-phased using SHAPEIT4 (v4.1.3)^65^, then imputed using Minimac4^66^ with African Genome Resources reference panel by Sanger Institute and 1000 Genomes Project phase3 as reference. Single nucleotide variants with imputation score <0.8, or HWE p-value ≤1×10^−6^, or minor allele frequency (MAF) lower than the threshold set in each ancestral group based upon their sample size (EA, 0.0005; AA, 0.001; LA, 0.005; EAA, 0.01; SAA, 0.01) were removed before association analysis.

Participants with at least one inpatient or two outpatient International Classification of Diseases (ICD)-9/10 codes for AUD were assigned as AUD cases, while participants with zero ICD codes for AUD were controls. Those with one outpatient diagnosis were excluded from the analysis. In total, 80,028; 36,330; 10,150; 701; and 107 cases were included in EUR, AFR, LA, EAS, and SAS, respectively; 368,113; 79,100; 28,812; 6,254; and 389 controls were included in EUR, AFR, LA, EAS, and SAS, respectively. BOLT-LMM^67^ was used to correct for relatedness, with age, sex, and the first 10 PCs as covariates.

### UK Biobank (UKB)

UKB released genotype and imputed data for ∼500,000 individuals from across the United Kingdom^17^ which were accessed through application 41910. UKB defined White-British (WB) participants genetically. For the non-WB individuals, we used PCA to classify them into different genetic groups as for MVP. Subjects with available AUDIT-P score were included in this study. The final sample included 132,001 WB (hereafter called UKB-EUR1) and 17,898 non-WB Europeans (hereafter called UKB-EUR2), and 1,220 SAS. SNPs with genotype call rate >0.95, HWE p-value >1×10^−6^, imputation score ≥0.8 and MAF ≥0.001 in EUR1 and EUR2 and ≥0.01 in SAS were kept for GWAS, BOLT-LMM was used for association correcting for relatedness, age, sex, and the first 10 PCs.

### FinnGen

Summary statistics for AUD from FinnGen data freeze 5 were downloaded from the FinnGen website (http://r5.finngen.fi/). Details of the genotyping, imputation and quality control for FinnGen data were described previously^20^. There were 8,866 AUD cases defined by ICD-8/9/10 codes and 209,926 controls. Association analysis was performed using SAIGE^68^ mixed-model with age, sex and 10 PCs as covariates. Positions of the variants were lifted over to build 37 (GRCh37/hg19) for meta-analysis.

### iPSYCH

The iPSYCH^21,22^ samples were selected from a baseline birth cohort comprising all singletons born in Denmark between May 1, 1981, and December 31, 2008. The iPSYCH study was approved by the Scientific Ethics Committee in the Central Denmark Region (Case No 1-10-72-287-12) and the Danish Data Protection Agency.

AUD was diagnosed according to the ICD-10 criteria (F10.1 – F10.9 diagnosis codes). The iPSYCH cohort was established to investigate genetic risk for major psychiatric disorders (i.e., attention-deficit/hyperactivity disorder, schizophrenia, bipolar disorder, major depressive disorder, autism spectrum disorder) but not AUD (or PAU), so comorbidity of psychiatric disorders among these AUD cases is higher than expected for cases selected randomly from the population. Therefore, we generated a control group around five times as large as the case groups, and to correct for the bias introduced by high comorbidity of psychiatric disorders among cases, we included within the control group individuals with the above listed psychiatric disorders (without comorbid AUD) at a proportion equal to what was observed among the cases.

The samples were genotyped in two genotyping rounds referred to as iPSYCH1 and iPSYCH2. iPSYCH1 samples were genotyped using Illumina’s PsychChip array and iPSYCH2 samples using Illumina’s GSA v.2 (Illumina, San Diego, CA, USA). Quality control and GWAS were performed using the Ricopili pipeline^69^. More details can be found in ref. 70. GWAS were performed separately for iPSYCH1 (2,117 cases and 13,238 controls) and iPSYCH2 (1,024 cases and 5,732 controls) using dosages for imputed genotypes and additive logistic regression with the first 5 PCs (from the final PCAs) as covariates using PLINK v1.9^71^. Only variants with a MAF >0.01 and imputation score >0.8 were included in the final summary statistics.

### Queensland Berghofer Medical Research Institute (QIMR) cohorts

The Australian Genetics of Depression Study (AGDS) recruited >20,000 participants with major depression between 2017 and 2020. Recruitment and subject characteristics have been reported^24^: Participants completed an online self-report questionnaire. Lifetime AUD was assessed on DSM-5 criteria using the Composite International Diagnostic Interview (CIDI). A total of 6,726 subjects with and 4,467 without AUD were included in the present study.

The Australian twin-family study of alcohol use disorder (TWINS, including Australian Alcohol and Nicotine Studies) participants were recruited from adult twins and their relatives who had participated in questionnaire- and interview-based studies on alcohol and nicotine use and alcohol-related events or symptoms (as described in Heath et al.^72^). They were predominantly of EUR ancestry. Young adult twins and their non-twin siblings were participants in the Nineteen and Up study (19Up)^25^. 2,772 cases and 5,630 controls were defined using DSM-III-R and DSM-IV criteria. Most alcohol-dependent cases were mild, with 70% of those meeting alcohol dependence criteria reporting only three or four dependence symptoms and ≤5% reporting seven dependence symptoms.

The Australian Genetics of Bipolar Disorder Study (GBP) recruited >5,000 participants living with bipolar disorder between 2018 and 2021. The sample’s recruitment and characteristics have been reported^23^: Participants completed an online self-report questionnaire. Lifetime DSM-5 AUD was assessed using the CIDI.

Genotyping of QIMR cohorts was performed using Illumina Global Screening Array v2. Pre-imputation QC removed variants with GenTrain score <0.6, MAF <0.01, SNP call rate <95%, and Hardy-Weinberg equilibrium deviation (*p*<1×10^−6^). Variants were then imputed using the Michigan Imputation Server with the Haplotype Reference Consortium reference panel^66^. Association analysis was performed using SAIGE and the LOCO=TRUE flag with age, sex, 10 PCs and two imputation variables as covariates. Participants of non-EUR ancestry (defined as >6 standard deviations from the PC1 and PC2 centroids) were excluded. Association analyses were limited to variants with a MAF≥0.0001, MAC≥5, and an R^2^≥0.1.

### Psychiatric Genomics Consortium (PGC)

Lifetime DSM-IV diagnosis of AD in both EUR and AFR ancestries were analyzed by PGC, with details reported previously^5^. This included 5,638 individuals from Australia. To avoid overlap with the new QIMR cohorts, we re-analyzed the PGC data without two Australian cohorts: Australian Alcohol and Nicotine Studies and Brisbane Longitudinal Twin Study. This yielded 9,938 cases and 30,992 controls of EUR ancestry and 3,335 cases and 2,945 controls of AFR ancestry.

### Yale-Penn 3

There are 3 phases of the Yale-Penn study defined by genotyping epoch; the first two were incorporated in the PGC study, thus they are included in the meta-analyses. Here, we included Yale-Penn 3 subjects as a separate sample. Lifetime AD was diagnosed based on DSM-IV criteria. Genotyping was performed in the Gelernter laboratory at Yale using the Illumina Multi-Ethnic Global Array, then imputed using Michigan Imputation Server with Haplotype Reference Consortium reference. We did PCA analyses to classify EAs (567 cases and 1,074 controls) and AAs (451 cases and 410 controls). Variants with MAF >0.01, HWE p-value >1×10^−6^ and imputation INFO score ≥0.8 were retained for association analyses using linear mixed models implemented in GEMMA^73^ and corrected for age, sex and 10 PCs.

### East Asian cohorts

Summary statistics for AUD/AD GWAS from 5 EAS cohorts (MVP EAS, Han Chinese–GSA, Thai METH–MEGA, Thai METH–GSA and Han Chinese–Cyto) were included in the cross-ancestry meta-analysis. Analyses of these five cohorts were previously published and the detailed QC can be found in ref. 27.

### Meta-analyses

Meta-analyses were performed using METAL^74^ with effective sample size weighting. For all the case-control samples, we calculated effective sample size as:

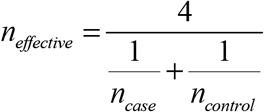

For AUDIT-P in UKB, a continuous trait, we used actual sample sizes for meta-analysis. For all meta-analyses within or across ancestries, variants with a heterogeneity test p-value <5×10^−8^ and variants with effective sample size <15% of the total effective sample size were removed. For the cross-ancestry and EUR within-ancestry meta-analyses, we required that variants were present in at least two cohorts. For the AFR and SAS within-ancestry meta-analyses, which are small samples, this was not required.

### Independent variants and conditional analyses

We identified the lead variants using PLINK with parameters of clumping region 500 kb and LD *r*^2^ 0.1. We then ran conditional analyses using GCTA-COJO^75^ to define conditionally independent variants among the lead variants using the 1000 Genomes Project phase3 as the LD reference panel. Any two independent variants <1 Mb apart whose clumped regions overlapped were merged into one locus.

### Cross-ancestry lookup

For the 85 independent variants associated in EUR, we looked up the associations in non-EUR groups. If the variants were not observed in another ancestry, we substituted proxy SNPs defined as associated with PAU (*p*<5×10^−8^) and in high LD with the EUR lead SNP (*r*^2^≥0.8).

### SNP-based heritability (*h*^2^)

SNP-based *h*^2^ for common SNPs mapped to HapMap3^76^ was estimated in EUR, AFR and LA ancestries using LDSC^77^; corresponding populations in the 1000 Genomes Project phase3 were used as LD reference panels. For PAU in EUR, we only estimated the observed-scale *h*^2^. For AUD, both observed-scale *h*^2^ and liability-scale *h*^2^ were estimated, using population lifetime prevalence estimates of 0.326, 0.220, and 0.229 in EUR, AFR, and LA, respectively^1^. These prevalence estimates were for lifetime DSM-5 AUD in the United States, which could introduce bias given the different definitions and prevalence in different cohorts. By default, LDSC removes SNPs with sample size <90^th^ percentile N/2. Here we skipped this filtering and kept all SNPs for analyses because we did basic filtering based on the number of cohorts and sample size. The final number of SNPs in the analyses range from 527,994 to 1.17 M.

### Cross-ancestry genetic correlation

We estimated the genetic correlations between different ancestries using Popcorn^78^, which can estimate both the genetic-effect correlation (ρ_ge_) as correlation coefficient of the per-allele SNP effect sizes and the genetic-impact correlation (ρ_gi_) as correlation coefficient of the ancestry-specific allele-variance-normalized SNP effect sizes. Populations in 1000 Genomes were used as reference for their corresponding population. A large sample size and number of SNPs are required for accurate estimation^78^, which explains the non-robust estimates for EAS and SAS samples.

### Within- and cross-ancestry fine-mapping

We did fine-mapping using MsCAVIAR^79^, which can leverage LD information from multiple ancestries to improve fine-mapping of causal variants. To reduce bias introduced by populations with small sample size, here we performed fine-mapping using summary statistics from the EUR, AFR and LA populations. Three sets of analyses were conducted. The first is within-ancestry fine-mapping for the 85 regions with independent variants in EUR using EUR summary data and 1000 Genomes Project phase3 EUR LD reference data. For each region, we selected SNPs which clumped (within 500 kb and LD *r*^2^>0.1) with the lead SNP and with *p*<0.05 for fine-mapping. We then calculated the pair-wise LD among the selected SNPs. If two SNPs were in perfect LD (*r*^2^=1, indicating that they are likely to be inherited together), we randomly removed one from the analysis. The second is cross-ancestry fine-mapping for the 100 regions with independent variants identified in cross-ancestry meta-analyses. For each region, we performed clumping (within 500 kb and LD *r*^2^>0.1) in EUR, AFR, and LA summary data for the lead SNP separately, to select 3 sets of SNPs (*p*<0.05) for fine-mapping, corresponding LD reference panels from 1000 Genomes Project were used. For each set of SNPs, we calculated the pair-wise LD and randomly removed one SNP if *r*^2^=1. If the lead SNP was not presented in the EUR SNP set, we did not perform fine-mapping for this region. Loci with limited numbers of variants cannot have convergent results, so they are not included in the results. After that, this cross-ancestry analysis included 92 regions. For the 10 regions in which the lead SNPs are missing in both AFR and LA populations, we did within-ancestry fine-mapping in EUR instead to keep the lead SNP (cross-ancestry fine-mapping will only analyze the SNPs common in analyzed ancestries). Third, because the credible set length identified is related to the number of variants in the input, to provide a more direct comparison between the cross-ancestry fine-mapping and the fine-mapping using information only from EUR, we used the same lists of SNPs from the above 92 regions in the cross-ancestry fine-mapping as for the EUR-only fine-mapping. “Credible set” was defined as plausible causal variants with accumulated posterior inclusion probability (PIP) > 99%. For each credible set, we report the variant with the highest PIP. We assumed that each locus contains only one causal variant by default, and increased to three at maximum if the analysis unable to converge.

### Gene-based association analyses

We performed gene-based association analysis for PAU or AUD in multiple ancestries using MAGMA implemented in FUMA^32,33^. Default settings were applied. Bonferroni corrections for the number of genes tested (range from 18,390 to 19,002 in different ancestries) were used to determine genome-wide significant genes.

### Transcriptome-wide association study (TWAS)

For PAU in EUR, we performed TWAS using S-PrediXcan to integrate transcriptomic data from GTEx. With prior knowledge that PAU is a brain-related disorder (evidenced by significant enrichment of gene expression in several brain tissues)^6^, 13 brain tissues were analyzed. The transcriptome prediction model database and the covariance matrices of the SNPs within each gene model were downloaded from the PredictDB repository (http://predictdb.org/). Significance of the gene-tissue association was determined following Bonferroni correction for the total number of gene-tissue pairs (*p*<0.05/166,064=3.01×10^−7^). We also used S-MultiXcan^36^ to integrate evidence across the 13 brain tissues using multivariate regression to improve association detection. In total, 18,383 genes were tested in S-MultiXcan, leading to a significance p-value threshold of 2.72×10^−6^.

### Association with chromatin interactions in brain

We used H-MAGMA^37^, a computational tool that incorporates brain chromatin interaction profiles from Hi-C, to identify risk genes associated with PAU based on EUR inputs. Six brain annotations were used: fetal brain, adult brain, adult midbrain dopaminergic, iPSC-derived astrocyte, iPSC-derived neuron and cortical neuron. In total, 319,903 gene-chromatin associations were analyzed across the six brain annotations. Significant genes were those with a p-value below the Bonferroni corrected value for the total number of tests (*p*<0.05/319,903=1.56×10^−7^).

### Probabilistic fine-mapping of TWAS

We did fine-mapping for TWAS in EUR using FOCUS^38^, a method that models correlation among TWAS signals to assign a PIP for every gene in the risk region to explain the observed association signal. The estimated credible set containing the causal gene can be prioritized for functional assays. FOCUS used 1000 Genomes Project EUR samples as the LD reference and multiple eQTL reference panel weights that include GTEx_v7^80^, The Metabolic Syndrome in Men^81^, Netherlands Twin Register^82^, Young Finns Study^83^, and CommonMind Consortium^84^. Under the model of PAU as substantially a brain disorder, we did fine-mapping while prioritizing predictive models using a brain tissue-prioritized approach.

### Drug repurposing

To match inferred transcriptional patterns of PAU with transcriptional patterns induced by perturbagens, we related our S-PrediXcan results to signatures from the Library of Integrated Network-based Cellular Signatures (LINCs) L1000 database^85^. This database catalogues *in vitro* gene expression profiles (signatures) from thousands of compounds >80 human cell lines (level 5 data from phase I: GSE92742 and phase II: GSE70138). Our analyses included signatures of 829 chemical compounds in five neuronal cell-lines (NEU, NPC, MNEU.E, NPC.CAS9 and NPC.TAK). To test significance of the association between PAU signatures and LINCs perturbagen signatures we followed the procedure from So et al^86^. Briefly, we computed weighted (by proportion of heritability explained) Pearson correlations between transcriptome-wide brain associations and *in vitro* L1000 compound signatures using the *metafor* package^87^ in R. We treated each L1000 compound as a fixed effect incorporating the effect size (*r*_weighted_) and sampling variability (se^2^) from all signatures of a compound (e.g., across all time points and doses). We only report those perturbagens that were associated after Bonferroni correction (*p*<0.05/829=6.03×10^−5^).

### Cross-ancestry polygenic risk score

We used PRS-CSx^40^, a method that couples genetic effects and LD across ancestries via a shared continuous shrinkage prior, to calculate the posterior effect sizes for SNPs mapped to HapMap3. Three sets of AUD GWAS summary data were use as input and corresponding posterior effect sizes in each ancestry were generated: EUR (without AUDIT-P from UKB, N_effective_=352,373), AFR (N_effective_=105,433), and LA (N_effective_=30,023). Three sets of AUD PRS based on the posterior effect sizes were calculated for UKB-EUR1 and UKB-EUR2 individuals using PLINK, following standardization (zero mean and unit variance) for each PRS. For each related pair (≥3^rd^-degree, kinship coefficient ≥0.0442 as calculated by UKB), we removed the subject with the lower AUDIT-P score, or randomly if they had the same score, leaving 123,565 individuals in UKB-EUR1 and 17,401 in UKB-EUR2. Then we ran linear regression for AUDIT-P in UKB-EUR2 as a validation dataset using PRS_EUR_, PRS_AFR_ and PRS_LA_ as independent variables. The corresponding regression coefficients were used as weights in the test dataset (UKB-EUR1) to calculate the final PRS: PRS_final_ = ω_EUR_*PRS_EUR_ + ω_AFR_*PRS_AFR_ + ω_LA_*PRS_LA_. We used linear regression to test the association between AUDIT-P and PRS_final_ after standardization, correcting for age, sex, and the first 10 PCs. We also ran a null model of association between AUDIT-P and covariates only, to calculate the variance explained (R^2^) by PRS_final_. For comparison, we also calculated PRS in UKB-EUR1 using only the AUD summary data in EUR using PRS-CS^88^, then calculated the variance explained by PRS_single_. The improved PRS association was measured as the difference of the variance explained (ΔR^2^).

### Genetic correlation

Genetic correlations (*r*_g_) between PAU or AUD and traits of interest were estimated using LDSC^89^. For EUR, we tested *r*_g_ between PAU and 49 traits using published summary data and the EUR LD reference from the 1000 Genomes Project. *r*_g_s with p-value <1.02×10^−3^ were considered significant. For AFR, we tested *r*_g_ between AUD and 13 published traits in AFR using MVP in-sample LD (most of the analyzed AFR were from MVP) built from 1000 randomly-selected AFR subjects by cov-LDSC^90^. *r*_g_s with p-value <3.85×10^−3^ (0.05/13) in AFR were considered as significant. For comparison, we also tested *r*_g_s using 1000 Genomes AFR as LD reference, which showed similar estimates.

### PAU PRS for phenome-wide associations

We calculated PRS using PRS-continuous shrinkage (PRS-CS) for PAU (in EUR) and AUD (in AFR) in four independent datasets [Vanderbilt University Medical Center’s Biobank (BioVU), Mount Sinai (Bio*Me™*), Mass General Brigham Biobank (MGBB)^91^ and Penn Medicine Biobank (PMBB)^92^] from the PsycheMERGE Network, followed by phenome-wide association studies. Details for each dataset are described below.

### BioVU

Genotyping of individuals was performed using the Illumina MEGEX array. Genotypes were filtered for SNP and individual call rates, sex discrepancies, and excessive heterozygosity using PLINK. Imputation was conducted using the Michigan Imputation Server based on the Haplotype Reference Consortium reference panel. PCA using FlashPCA2^93^ combined with CEU, YRI and CHB reference sets from the 1000 Genomes Project Phase 3 was conducted to determine participants of AFR and EUR ancestry. One individual from each pair of related individuals was removed (pi-hat>0.2). This resulted in 12,384 AFR and 66,903 EUR individuals for analysis.

#### BioMe*™*

The Bio*Me™* Biobank: The Illumina Global Screening Array was used to genotype the Bio*Me™* samples. The SNP-level quality control (QC) removed SNPs with (1) MAF <0.0001 (2) HWE p-value ≤1×10^−6^ and (3) call rate <98%. The individual-level QC removed participants with (1) sample call rate <98% and (2) heterozygosity F coefficient ≥3 standard deviations. In addition, one individual from each pair of related samples with a genomic relatedness (proportion IBD) >0.125 was removed (--rel-cutoff=0.125 in PLINK). Imputation was performed using 1000 Genomes Phase 3 data. Each ancestry was confirmed by the genetic PC plot. A final sample size of 4,727 AFR and 9,544 EUR individuals were included for this study.

### MGBB

Individuals in the Mass General Brigham Biobank (MGBB) were genotyped using the Illumina Multi-Ethnic Global array with hg19 coordinates. Variant-level quality control filters removed variants with a call rate <98% and those that were duplicated across batches, monomorphic, not confidently mapped to a genomic location, or associated with genotyping batch. Sample-level quality control filters removed individuals with a call rate less than 98%, excessive autosomal heterozygosity (±3 standard deviations from the mean), or discrepant self-reported and genetically inferred sex. PCs of ancestry were calculated in the 1000 Genomes Phase 3 reference panel and subsequently projected onto the MGBB dataset, where a Random Forest classifier was used to assign ancestral group membership for individuals with a prediction probability >90%. The Michigan Imputation Server was then used to impute missing genotypes with the Haplotype Reference Consortium dataset serving as the reference panel. Imputed genotype dosages were converted to hard-call format and subjected to further quality control, where SNPs were removed if they exhibited poor imputation quality (INFO<0.8), low minor allele frequency (<1%), deviations from Hardy-Weinberg equilibrium (*p*<1×10^−10^), or missingness (variant call rate <98%). Only unrelated individuals (pi-hat<0.2) of EUR ancestry were included in the present study. These procedures yielded a final analytic sample of 25,698 individuals in the MGBB.

### PMBB

Genotyping of individuals was performed using the Illumina Global Screening Array. Quality control removed SNPs with marker call rate <95% and sample call rate <90%, and individuals with sex discrepancies. Imputation was performed using Eagle2^94^ and Minimac4 on the TOPMed Imputation Server. One individual from each pair of related individuals (pi-hat threshold of 0.25) were removed from analysis. PCA was conducted using smartpca^62,63^ and the Hapmap3 dataset to determine genetic ancestry. This resulted in 10,383 AFR and 29,355 EUR individuals for analysis.

### PheWAS

The AFR AUD PRS and EUR PAU PRS scores in each dataset were standardized for the PheWAS analyses. International Classification of Diseases (ICD)-9 and -10 codes were extracted from the electronic health record and mapped to phecodes. Individuals were considered cases if they had two instances of the phecode. We conducted PheWAS by fitting a logistic regression for each phecode within each biobank. Covariates included sex, age and the top 10 PCs. PheWAS results were meta-analyzed within each ancestral group across biobanks (AFR=27,494, EUR=131,500) using the PheWAS package^95^ in R.

### Yale-Penn

Quality control and creation of the PheWAS dataset have been described previously^45^. We calculated PRS for PAU in EUR and AUD in AFR (using summary statistics that leave out the Yale-Penn 3 and PGC sample which includes Yale-Penn 1 and 2). We conducted PheWAS by fitting logistic regression models for binary traits and linear regression models for continuous traits. We used sex, age at recruitment, and the top 10 genetic PCs as covariates. We applied a Bonferroni correction to control for multiple comparisons.

## Data Availability

The full summary-level association data from the meta-analysis are available upon request to the corresponding authors and through dbGaP (accession number phs001672).

https://www.ncbi.nlm.nih.gov/projects/gap/cgi-bin/study.cgi?study_id=phs001672.v8.p1

## Acknowledgements

This research used data from the Million Veteran Program and was supported by funding from the Department of Veterans Affairs Office of Research and Development, Million Veteran Program Grant #I01CX001849, #I01BX004820, #I01BX003341, and the VA Cooperative Studies Program (CSP) study #575B, MVP004, and MVP025. This publication does not represent the views of the Department of Veterans Affairs or the United States Government. Supported also by NIH (NIAAA) P50 AA12870 (J.H.K.) and K01 AA028292 (R.L.K.), a NARSAD Young Investigator Grant 27835 from the Brain & Behavior Research Foundation (H.Z.), NCI R21 CA252916 (H.Z.), R01 AA026364 (J.G.), NIAAA T32 AA028259 (J.D.D.), NIMH R01 MH124839 (L.M.H.), NIAAA K01 AA030083 (A.S.H.), NIDA K01 DA051759 (E.C.J.), TRDRP (T29KT0526, T32IR5226, S.S.R.) and NIDA DP1 DA054394 (S.S.R.). This research used data from UK Biobank (project ID: 41910), a population-based sample of participants whose contributions we gratefully acknowledge. The data access is supported by Yale SCORE pilot grant (U54AA027989). We want to acknowledge the participants and investigators of the FinnGen study. D.D. was supported by the Novo Nordisk Foundation (NNF20OC0065561), the Lundbeck Foundation (R344-2020-1060). The iPSYCH team was supported by grants from the Lundbeck Foundation (R102-A9118, R155-2014-1724, and R248-2017-2003), NIH/NIMH (1U01MH109514-01 and 1R01MH124851-01 to A.D.B.) and the Universities and University Hospitals of Aarhus and Copenhagen. The Danish National Biobank resource was supported by the Novo Nordisk Foundation. High-performance computer capacity for handling and statistical analysis of iPSYCH data on the GenomeDK HPC facility was provided by the Center for Genomics and Personalized Medicine and the Centre for Integrative Sequencing, iSEQ, Aarhus University, Denmark (grant to A.D.B.). The Australian Genetics of Depression Study (AGDS) was primarily funded by the National Health and Medical Research Council (NHMRC) of Australia Grant No. 1086683 to N.G.M. N.G.M is supported by a NHMRC Investigator Grant (No. APP 1172990). We are indebted to all of the participants for giving their time to contribute to this study. We wish to thank all the people who helped in the conception, implementation, media campaign and data cleaning. We thank Richard Parker, Simone Cross, and Lenore Sullivan for their valuable work coordinating all the administrative and operational aspects of the AGDS project. We would also like to thank the research participants for making this work possible. The Australian Genetics of Bipolar Disorder Study (GBP) data collection was funded and data analysis was supported by the Australian NHMRC (No. APP1138514) to S.E.M. S.E.M. is supported by a NHMRC Investigator Grant (No. APP1172917). We thank the participants for giving their time and support for this project. We acknowledge and thank M. Steffens for her generous donations and fundraising support. The NHMRC (APP10499110) and the NIH (K99R00, R00DA023549) funded the 19Up study. Genotyping was funded by the NHMRC (389891). We thank the twins and their families for their willing participation in our studies. Funding for the Australian adult twin studies in which information on alcohol use and smoking status was obtained came from the US NIH (AA07535, AA07728, AA11998, AA13320, AA13321, AA14041, AA17688, DA012854 and DA019951); the Australian NHMRC (241944, 339462, 389927, 389875, 389891, 389892, 389938, 442915, 442981, 496739, 552485 and 552498); and the Australian Research Council (A7960034, A79906588, A79801419, DP0770096, DP0212016 and DP0343921). We acknowledge the work over many years of staff of the Genetic Epidemiology group at QIMR Berghofer Medical Research Institute (formerly the Queensland Institute of Medical Research) in managing the studies which generated the data used in this analysis. We also acknowledge and appreciate the willingness of study participants to complete multiple, and sometimes lengthy, questionnaires and interviews. Many of the participants were contacted originally through the Australian Twin Registry. This research also used summary data from the Psychiatric Genomics Consortium (PGC) Substance Use Disorders (SUD) working group. The PGC-SUD is supported by NIH grant R01DA054869. PGC-SUD gratefully acknowledges its contributing studies and the participants in those studies, without whom this effort would not be possible. We acknowledge the Penn Medicine BioBank (PMBB) for providing data and thank the patient-participants of Penn Medicine who consented to participate in this research program. We would also like to thank the PMBB team and Regeneron Genetics Center for providing genetic variant data for analysis. The PMBB is approved under IRB protocol# 813913 and supported by Perelman School of Medicine at University of Pennsylvania, a gift from the Smilow family, and the National Center for Advancing Translational Sciences of the NIH under CTSA award number UL1TR001878. This study was supported in part through the resources and staff expertise provided by the Charles Bronfman Institute for Personalized Medicine and The Bio*Me™* Biobank Program at the Icahn School of Medicine at Mount Sinai. Research reported in this paper was supported by the Office of Research Infrastructure of the NIH under award numbers S10OD018522 and S10OD026880. The content is solely the responsibility of the authors and does not necessarily represent the official views of the NIH.

