## Supplementary Figures 1-5 for "Multi-ancestry study of the genetics of problematic alcohol use in >1 million individuals"

**Supplementary Figure 1. Manhattan and QQ plots for PAU/AUD meta-analyses in different ancestries.** a, PAU meta-analysis in European ancestry. b, AUD meta-analysis in African ancestry. c, AUD in Latin Americans from MVP. d, PAU meta-analysis in South Asian ancestry.

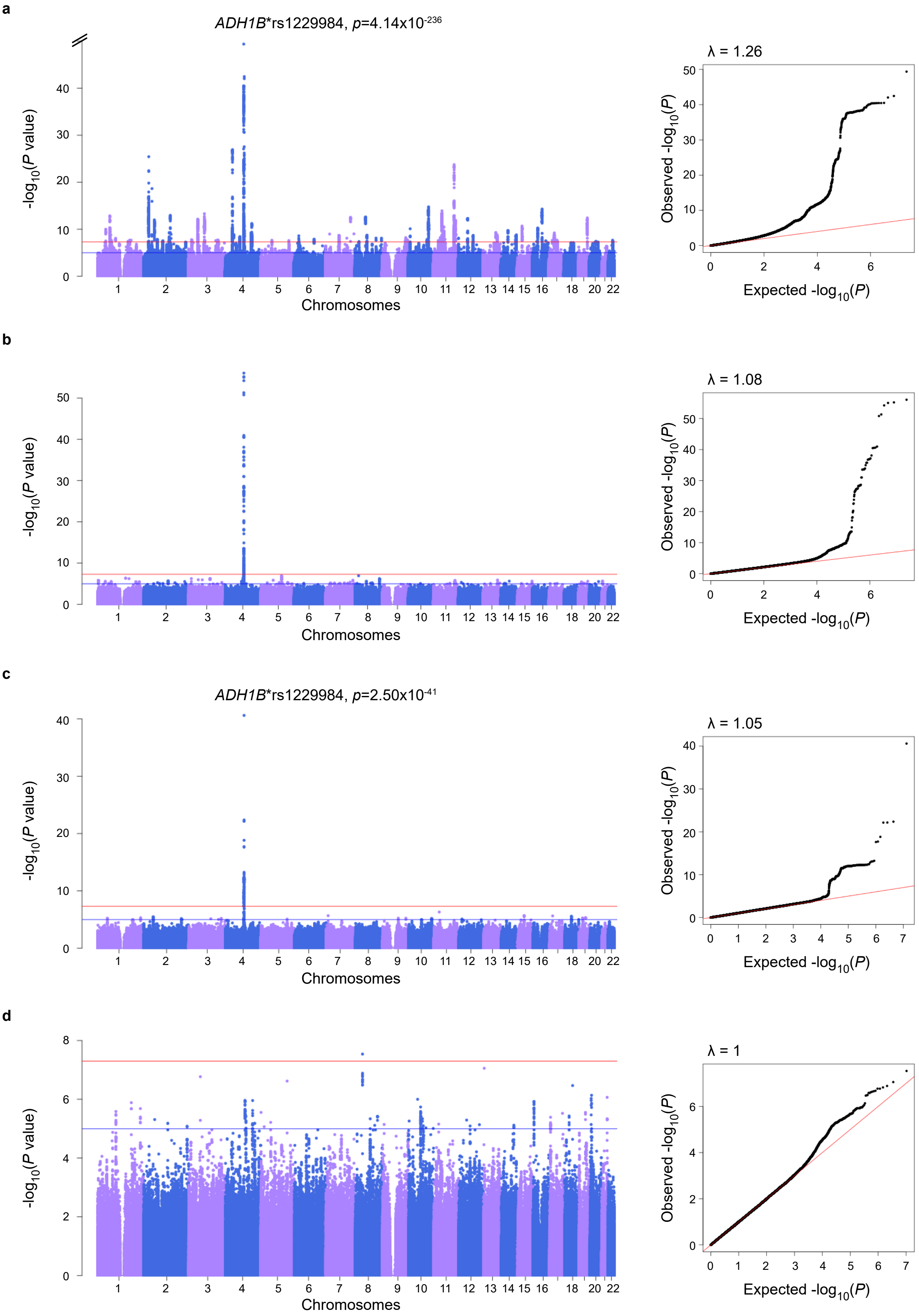

**Supplementary Figure 5. Phenome-wide associations with PAU PRS in PsycheMERGE EUR samples.**

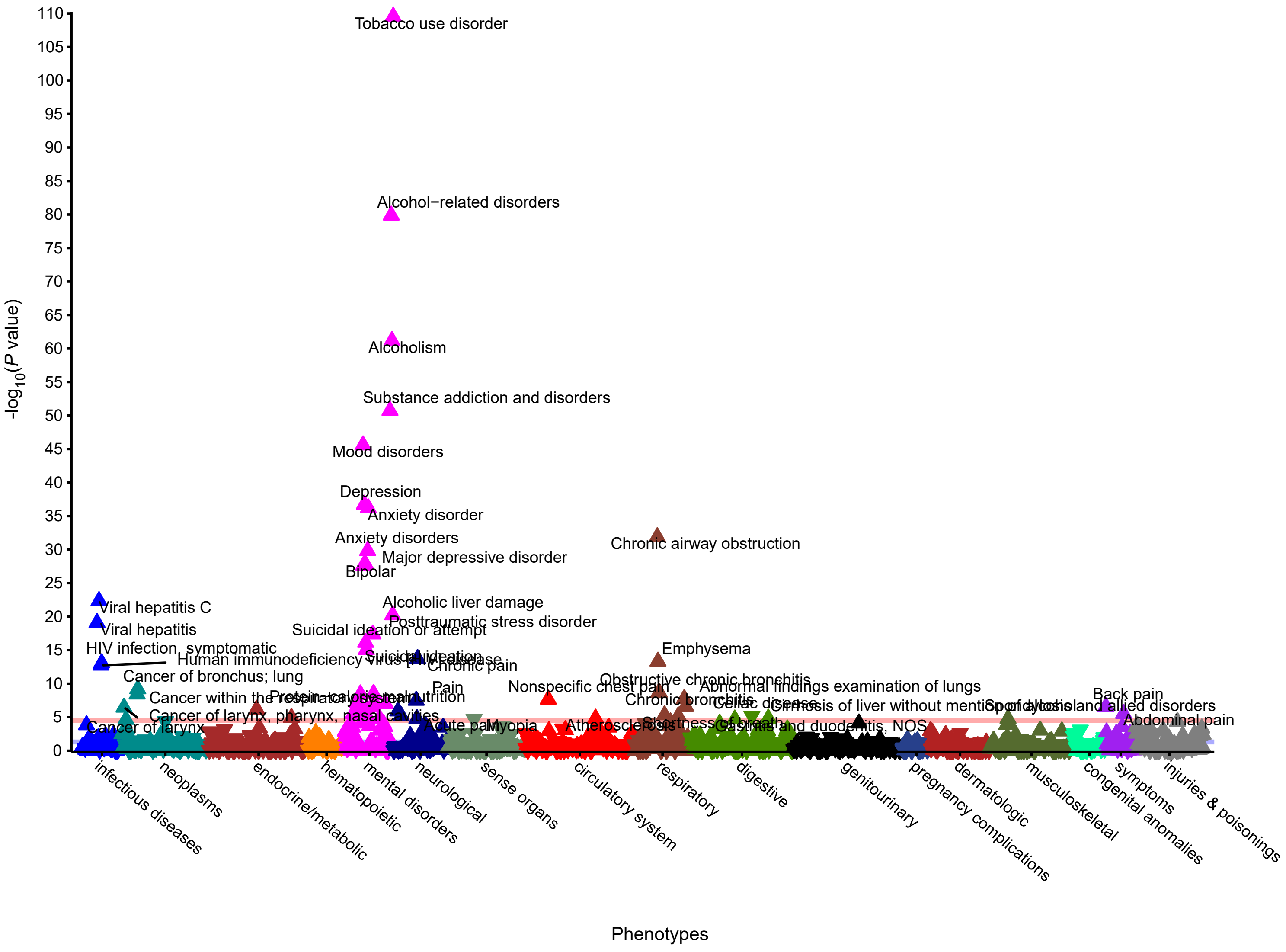

**Supplementary Figure 6. Phenome-wide associations with AUD PRS in PsycheMERGE AFR samples.**

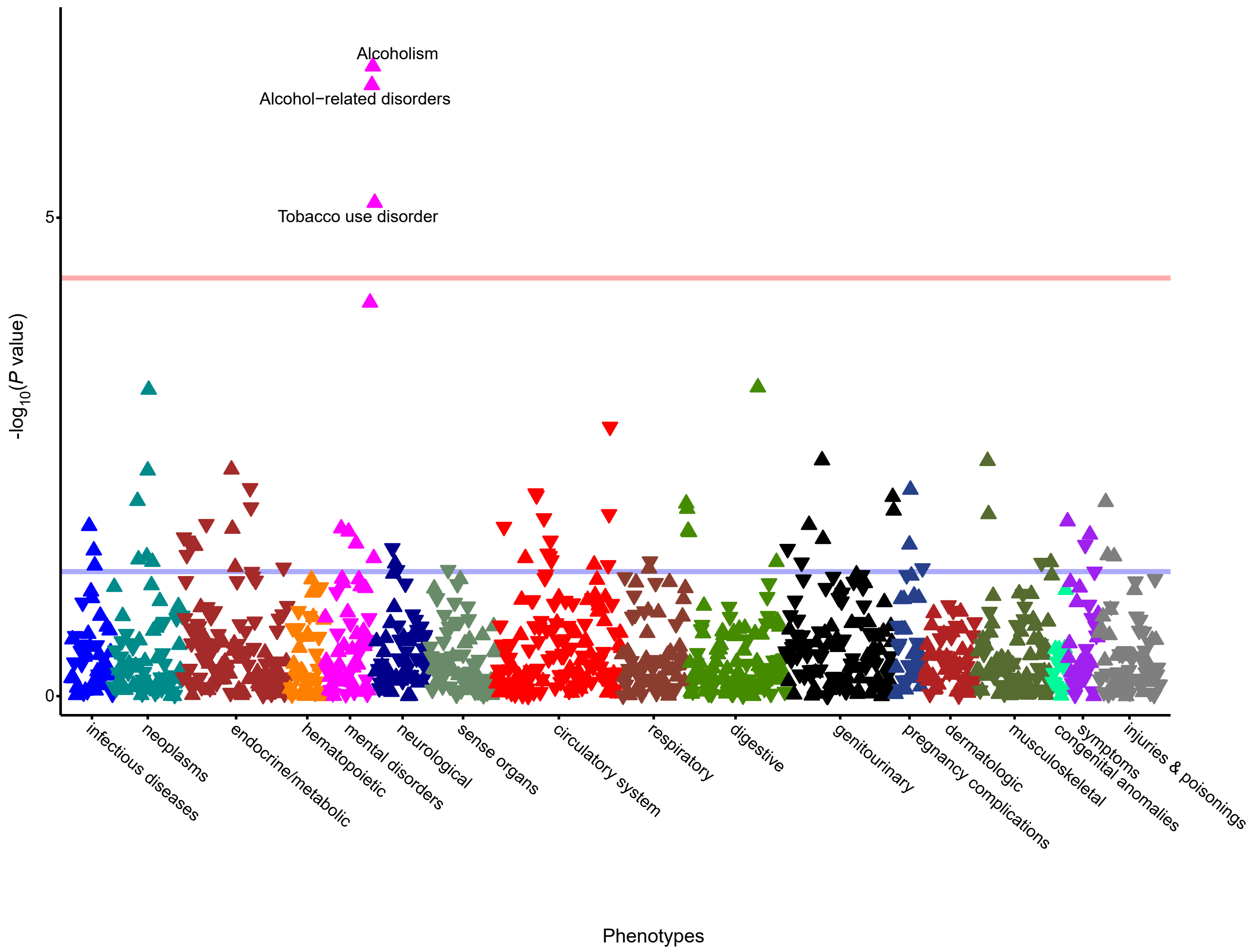

**Supplementary Figure 7. Phenome-wide associations with PAU PRS in Yale-Penn EUR samples.**

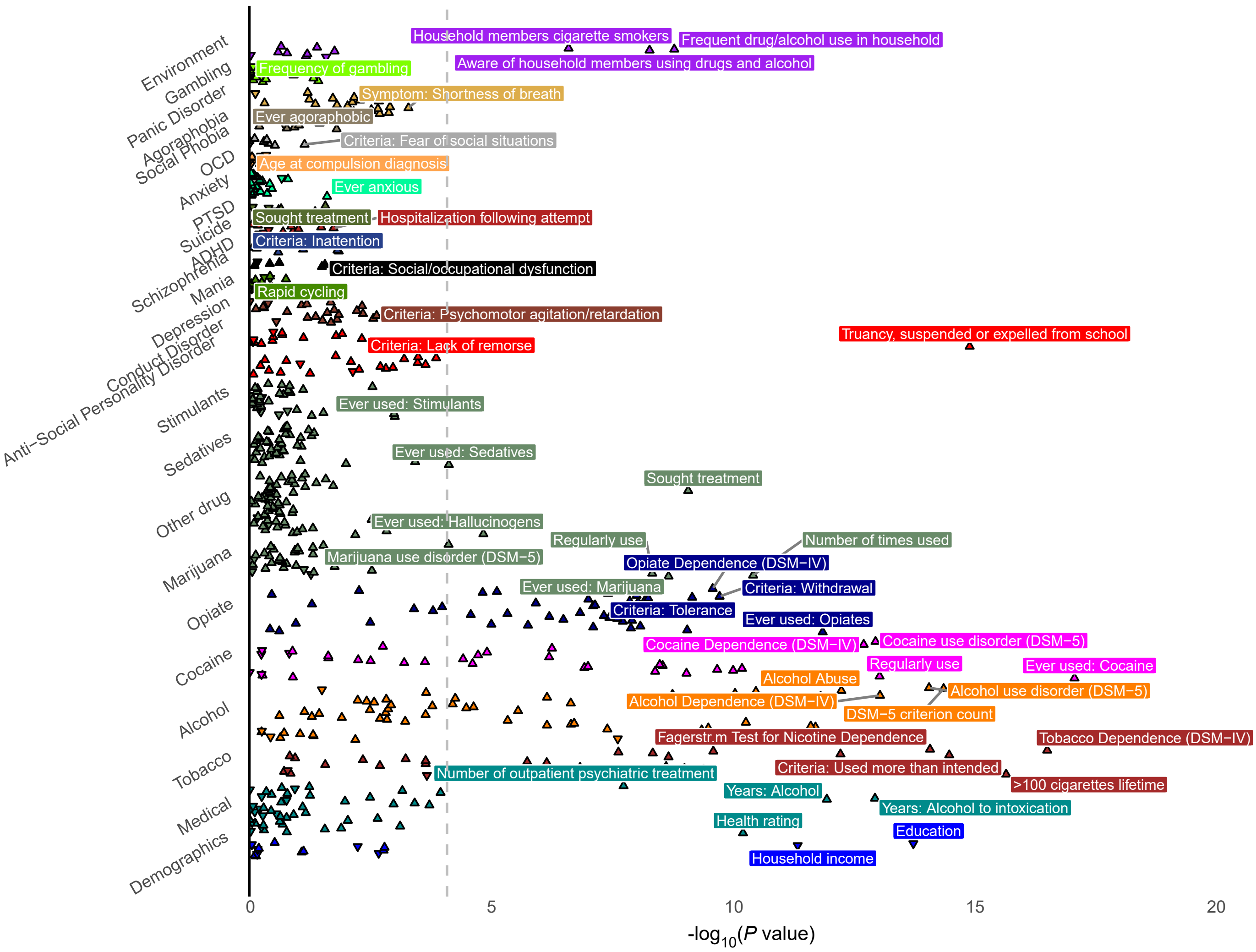

**Supplementary Figure 8. Phenome-wide associations with AUD PRS in Yale-Penn AFR samples.**

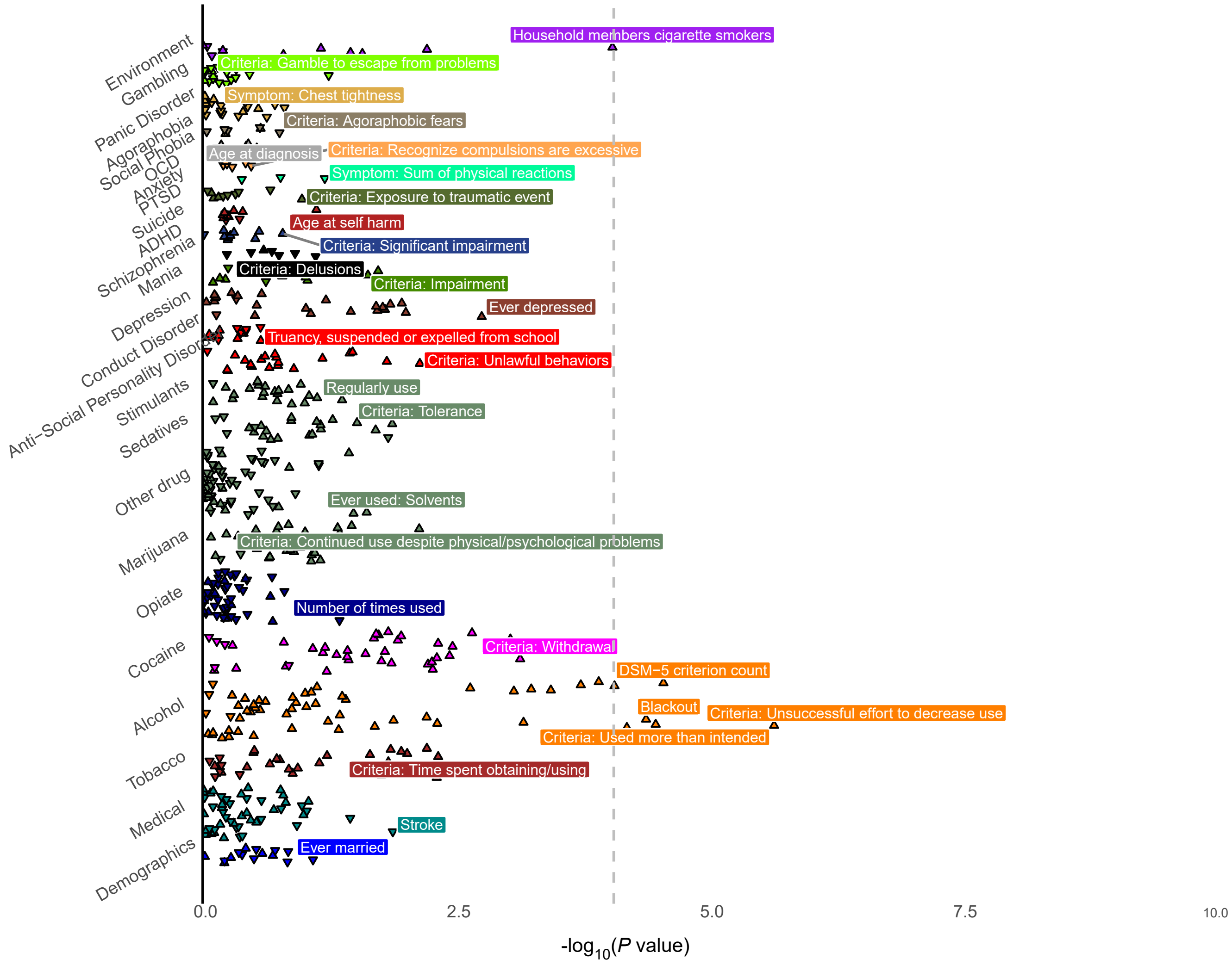
